# Latent classes of men’s intimate partner violence perpetration and attitudes towards gender norms: a UN multi-country, cross-sectional study in Asia and the Pacific

**DOI:** 10.1101/2022.02.07.22270580

**Authors:** Tiara C. Willie, Marina Katague, Nafisa Halim, Jhumka Gupta

## Abstract

**Objective:** To examine distinct patterns of IPV perpetration and examined gender equitable attitudes as a predictor of these patterns among men from six countries in Asia and the Pacific.

**Design:** 2011-12 UN Multi-country Study on Men and Violence cross-sectional study.

**Setting:** Households in Bangladesh, Cambodia, China, Indonesia, Sri Lanka and Papua New Guinea.

**Participants:** 10,178 men aged 18-49 years residing in Bangladesh, Cambodia, China, Indonesia, Sri Lanka and Papua New Guinea.

**Primary outcomes measure:** Our primary outcome was distinct patterns of IPV perpetration which were derived from multilevel latent class analyses.

**Results:** The odds of being assigned to the Low All Forms of IPV Perpetration class than the High All Forms of IPV Perpetration class was lower for men in the middle tertile group than men in the high tertile group for gender equitable attitudes. The odds of being assigned to the High Emotional IPV Perpetration class than the High All Forms of IPV Perpetration class was greater for men in the low tertile group than men in the high tertile group for gender equitable attitudes. The odds of being assigned to the High Physical/Emotional/Economic IPV Perpetration class than the High All Forms of IPV Perpetration class was lower for men in the low tertile group than men in the high tertile group for gender equitable attitudes.

**Conclusions:** Gender transformative interventions that use an adaptive, personalized approach to men’s typology of IPV perpetration may be beneficial to reduce violence against for women in the Asia-Pacific region.

**Strengths and limitations of the study:**

- This is one of the first studies to examined distinct patterns of intimate partner violence (IPV) perpetration among men in low- and middle-income countries, include the Asia-Pacific region.
- This study used a large sample of 10,178 men aged 18-49 years in the Asia-Pacific region.
- The measures of IPV perpetration were well-defined but the data was self-reported data, and IPV perpetration may be under-reported, if perceived as a private matter.
- The current analysis is a cross-sectional study and thus unable to make inferences regarding causality.

## INTRODUCTION

Multisectoral efforts are needed to respond to intimate partner violence (IPV) as a human rights violation and serious threat to global public health,^1^ as women are more at risk of experiencing violence in intimate relationships than experiencing violence in any other context.^2^ One in three women are estimated to experience IPV over their lifetime.^3^ Common underlying factors that contribute to the burden of IPV include male feelings of entitlement to forced sex within marriage.^4,5^ Intimate partners who perpetrate physical or sexual violence also tend to practice controlling behaviors,^2^ which all serve to threaten the health, social well-being, and economic growth of victims. IPV can result in adverse health outcomes,^6^ including mental health,^7^ physical health,^8^ and sexual health.^9,10^

Prevention research examining the different forms of IPV perpetration among men in Asia and the Pacific is further warranted. Southeast Asia has a high prevalence (37.7%) of physical and sexual IPV among ever-partnered women worldwide.^3^ In the countries included in the current study, gender-based violence behaviors, such as domestic violence and spousal abuse, are especially pervasive.^5,11^ For example, 75% of married Bangladeshi women in a nationally representative sample experienced physical and/or sexual violence.^12^

Despite the pervasive prevalence of IPV across the Asia-Pacific region, research seeking to understand male perpetration of different IPV behaviors in this region is relatively scarce. Prevalence of IPV is varied across the regions included in the study, with highest rates reported in Papua New Guinea.^5^ Additional research directly with men is needed to understand the underlying drivers of violent behaviors to transform the context which makes violence by men against women a societal norm. Although factors associated with men’s perpetration of violence against women varied across Bangladesh, Cambodia, China, Indonesia, Papua New Guinea, and Sri Lanka, IPV was found to be related to unequal gender norms and power inequalities.^5,13,14^ Indeed, despite a majority of respondents reporting they believed in gender equality across the included sites in Asia and the Pacific, rape was most commonly motivated by a sense of entitlement.^14^ Evidence suggests that men’s attitudes about gender norms, and their perpetuation through the social reproduction of these norms in institutions and culture, directly influence behavior,^15^ including the perpetration of physical and sexual violence.^16^

Further, violence is a form of discipline used to enforce gender norms.^17,18^ Gender-transformative approaches that engage men’s attitudes and behaviors have shown to be a promising approach to reduce IPV.^19^ Notably, existing intervention studies have had differential impacts on various forms of IPV.^1-4^ Investigating differences in men’s attitudes and how they relate to patterns of IPV perpetration may help inform how and for whom these approaches may be the most efficacious.

Identifying subgroups of men who perpetrate IPV to identify differing attitudes toward gender norms may help tailor intervention for violence prevention. Latent class analysis (LCA) is a person-centered approach used to identify subgroups based on patterns of behavior,^21,22^ which has recently been used to understand heterogeneity in IPV experiences globally.^23-26^ LCA may advance our understanding of IPV perpetration by investigating diversity of relationships between violence-related variables,^27^ as specific gender equitable norms may be driving specific patterns of IPV against women. Teasing apart underlying factors by behaviors may serve to inform nuanced approaches for programming in the future. To date, the authors are unaware of any study that has adopted this approach to examine IPV perpetration in the region of Asia and the Pacific or any other LMIC.

The current study uses data from the UN Multi-country Study on Men and Violence in Asia and the Pacific to conduct the following aims: (1) use LCA to identify distinct typologies of men based on their perpetration of intimate partner violence, and (2) examine gender equitable attitudes as correlates of men’s IPV perpetration typologies.

## MATERIALS AND METHODS

The methods below have been described elsewhere.^5-7^

### Study Procedures

The current analytic sample data were drawn from the UN Multi-country Study on Men and Violence, a cross-sectional, multicountry population-based quantitative household survey of 10,178 men and 3,106 women in nine sites across six countries (Bangladesh, Cambodia, China, Indonesia, Sri Lanka and Papua New Guinea). The study was a collaboration between the Partners for Prevention with the Medical Research Council of South Africa and country research teams. The survey was administered from 2011 to 2012. In every research site, a representative sample of men aged 18-49 were recruited using a multi-stage cluster sample strategy. Male participants were interviewed by male interviewers and data was collected using personal digital assistants. All research sites had a response rate of 70%, except in Sri Lanka, which had a response rate of 59%. The study followed international ethical and safety standards for research on violence against women.^8^ In the original study, participants were asked to sign consent forms if they felt comfortable or give verbal consent to participate.

The present study was exempted by [Institution masked for peer review] IRB Number: 1503280-1 because the analyses were secondary, and data were de-identified. Given our focus on men’s perpetration of IPV, we restricted our analytic sample to male participants across all research sites (N=10,178).

### Patient and Public Involvement

Since the current study is a secondary data analysis of deidentified participant data, it was not appropriate or possible to involve patients or the public in the design, or conduct, or reporting, or dissemination plans of our research study.

### Measures

Latent classes were based on 18 behaviors related to lifetime. Men were asked a series of behavior-specific questions related to current or former IPV based on the WHO Multi-country Study on Women’s Health and Domestic Violence questionnaire for women,^9^ and a South African survey adapted for men.^10^ Men were asked questions related to perpetration of five acts of emotional abuse (e.g., belittling, insults), five acts of economic abuse (e.g., prohibited her employment), six acts of physical abuse (e.g., slapped or thrown something), and 2 acts of sexual abuse (e.g., forced sex). For each of the 18 items, an affirmative response (yes) was coded as perpetrating that specific form of IPV against a female.

Gender equitable attitudes was assessed using the Gender-Equitable Men (GEM) scale.^11^ Participants were asked 8 gender-related statements across five domains gender norms (e.g., “A man should have the final word about decisions in his home”), violence (e.g., “If someone insults me, I will defend my reputation with force if I have to”), sexuality (e.g., “Men need sex more than women do”), toxic masculinity (e.g., “To be a man, you need to be tough”) and reproductive health (e.g., “It’ is a woman’s responsibility to avoid getting pregnant”). Participants were asked to respond on a 4-point Likert scale ranging from Strongly Agree (1) to Strongly Disagree (4). Responses were summed to create a total score and the total score was categorized into tertiles to represent low, medium and high levels of gender equitable attitudes and beliefs. A total score for each domain was created as well. The tertiles variable was used in the main analyses and the domain-specific total scores were used in the post-hoc analyses. Creating scores based on tertiles^12,13^ and domains^11,14^ is consistent with previous research.

Men were asked to report socio-demographics including age, education, marital/cohabitating status, employment, and number of children.

### Data Analysis

Descriptive statistics (frequencies, means) were conducted to describe characteristics of the analytic sample. Latent class analyses were based on 18 indicators of IPV perpetration. Model fit was assessed based on the fit statistics: lowest values for adjusted Bayesian Information Criteria, Lo-Mendell-Rubin likelihood ratio *p*-value<.05, sample size and epidemiologically relevant.^15^ An entropy value greater than or equal to 0.80 was considered optimal.^16^ Latent classes were named based upon whether the probability of reporting the latent indicator for IPV perpetration was greater than or equal to 0.50. Finally, latent class regression was used to test associations between gender equitable attitudes and latent class membership while controlling for age, education, marital/cohabitating status, employment and whether they had children.

Odds ratios (ORs), 95% confidence intervals (CI), and p-values <0.05 were used to assess significance in latent class regression models. All analyses accounted for the multistage structure and clustering of sites within countries. Descriptive analyses were conducted using SAS 9.4. Latent class analyses were conducted using Mplus 7.0.^17^

## RESULTS

### Prevalence of Socio-demographics of Overall Sample

**Table 1** displays characteristics of the overall sample of male participants. The majority of male participants were 34 or younger: 26.3% were 18-24 years and 33.4% were 25-34 years of age and the mean number of children were two. Over two-thirds of the male participants had at least some high school education. The majority were employed (82.8%) and either married or cohabited (66.7%). On average, men had between one and two children across countries.

**Table 1.**
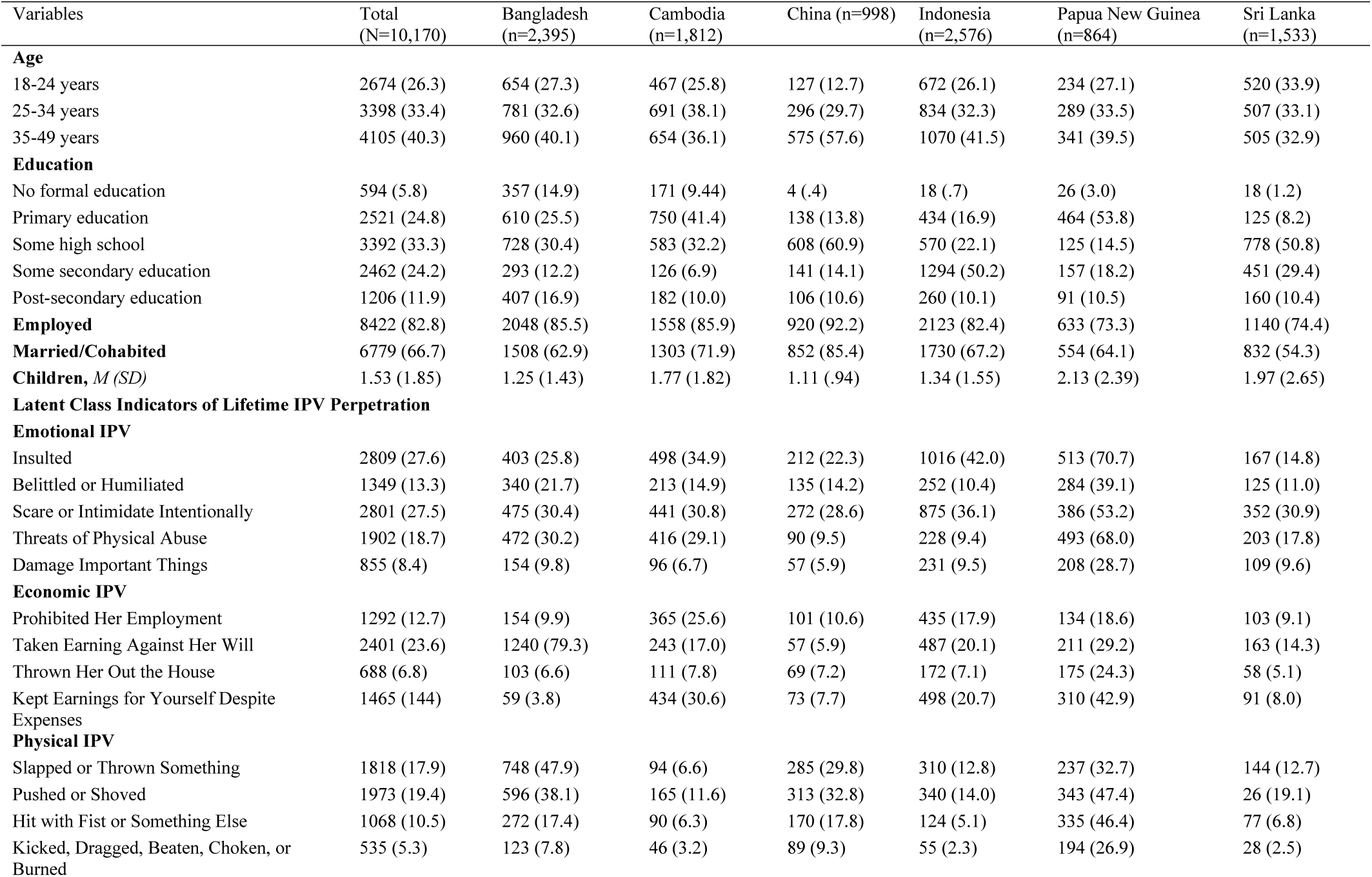
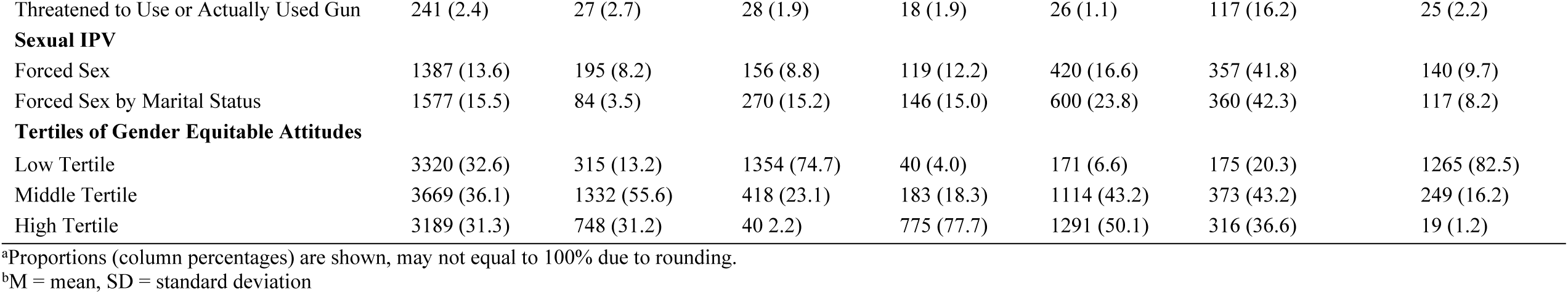
Prevalence of Demographics, Lifetime IPV Perpetration and Gender Equitable Attitudes Categories among Male Participants Recruited for UN Multi-Country Study on Men and Violence (total sample and by country)

| Variables | Total<br>(N=10,170) | Bangladesh<br>(n=2,395) | Cambodia<br>(n=1,812) | China (n=998) | Indonesia<br>(n=2,576) | Papua New Guinea<br>(n=864) | Sri Lanka<br>(n=1,533) |
| --- | --- | --- | --- | --- | --- | --- | --- |
| **Age** |  |  |  |  |  |  |  |
| 18-24 years | 2674 (26.3) | 654 (27.3) | 467 (25.8) | 127 (12.7) | 672 (26.1) | 234 (27.1) | 520 (33.9) |
| 25-34 years | 3398 (33.4) | 781 (32.6) | 691 (38.1) | 296 (29.7) | 834 (32.3) | 289 (33.5) | 507 (33.1) |
| 35-49 years | 4105 (40.3) | 960 (40.1) | 654 (36.1) | 575 (57.6) | 1070 (41.5) | 341 (39.5) | 505 (32.9) |
| **Education** |  |  |  |  |  |  |  |
| No formal education | 594 (5.8) | 357 (14.9) | 171 (9.44) | 4 (.4) | 18 (.7) | 26 (3.0) | 18 (1.2) |
| Primary education | 2521 (24.8) | 610 (25.5) | 750 (41.4) | 138 (13.8) | 434 (16.9) | 464 (53.8) | 125 (8.2) |
| Some high school | 3392 (33.3) | 728 (30.4) | 583 (32.2) | 608 (60.9) | 570 (22.1) | 125 (14.5) | 778 (50.8) |
| Some secondary education | 2462 (24.2) | 293 (12.2) | 126 (6.9) | 141 (14.1) | 1294 (50.2) | 157 (18.2) | 451 (29.4) |
| Post-secondary education | 1206 (11.9) | 407 (16.9) | 182 (10.0) | 106 (10.6) | 260 (10.1) | 91 (10.5) | 160 (10.4) |
| **Employed** | 8422 (82.8) | 2048 (85.5) | 1558 (85.9) | 920 (92.2) | 2123 (82.4) | 633 (73.3) | 1140 (74.4) |
| **Married/Cohabited** | 6779 (66.7) | 1508 (62.9) | 1303 (71.9) | 852 (85.4) | 1730 (67.2) | 554 (64.1) | 832 (54.3) |
| **Children, *M (SD)*** | 1.53 (1.85) | 1.25 (1.43) | 1.77 (1.82) | 1.11 (.94) | 1.34 (1.55) | 2.13 (2.39) | 1.97 (2.65) |
| **Latent Class Indicators of Lifetime IPV Perpetration** |  |  |  |  |  |  |  |
| **Emotional IPV** |  |  |  |  |  |  |  |
| Insulted | 2809 (27.6) | 403 (25.8) | 498 (34.9) | 212 (22.3) | 1016 (42.0) | 513 (70.7) | 167 (14.8) |
| Belittled or Humiliated | 1349 (13.3) | 340 (21.7) | 213 (14.9) | 135 (14.2) | 252 (10.4) | 284 (39.1) | 125 (11.0) |
| Scare or Intimidate Intentionally | 2801 (27.5) | 475 (30.4) | 441 (30.8) | 272 (28.6) | 875 (36.1) | 386 (53.2) | 352 (30.9) |
| Threats of Physical Abuse | 1902 (18.7) | 472 (30.2) | 416 (29.1) | 90 (9.5) | 228 (9.4) | 493 (68.0) | 203 (17.8) |
| Damage Important Things | 855 (8.4) | 154 (9.8) | 96 (6.7) | 57 (5.9) | 231 (9.5) | 208 (28.7) | 109 (9.6) |
| **Economic IPV** |  |  |  |  |  |  |  |
| Prohibited Her Employment | 1292 (12.7) | 154 (9.9) | 365 (25.6) | 101 (10.6) | 435 (17.9) | 134 (18.6) | 103 (9.1) |
| Taken Earning Against Her Will | 2401 (23.6) | 1240 (79.3) | 243 (17.0) | 57 (5.9) | 487 (20.1) | 211 (29.2) | 163 (14.3) |
| Thrown Her Out the House | 688 (6.8) | 103 (6.6) | 111 (7.8) | 69 (7.2) | 172 (7.1) | 175 (24.3) | 58 (5.1) |
| Kept Earnings for Yourself Despite Expenses | 1465 (14.4) | 59 (3.8) | 434 (30.6) | 73 (7.7) | 498 (20.7) | 310 (42.9) | 91 (8.0) |
| **Physical IPV** |  |  |  |  |  |  |  |
| Slapped or Thrown Something | 1818 (17.9) | 748 (47.9) | 94 (6.6) | 285 (29.8) | 310 (12.8) | 237 (32.7) | 144 (12.7) |
| Pushed or Shoved | 1973 (19.4) | 596 (38.1) | 165 (11.6) | 313 (32.8) | 340 (14.0) | 343 (47.4) | 26 (19.1) |
| Hit with Fist or Something Else | 1068 (10.5) | 272 (17.4) | 90 (6.3) | 170 (17.8) | 124 (5.1) | 335 (46.4) | 77 (6.8) |
| Kicked, Dragged, Beaten, Choken, or Burned | 535 (5.3) | 123 (7.8) | 46 (3.2) | 89 (9.3) | 55 (2.3) | 194 (26.9) | 28 (2.5) |
| Threatened to Use or Actually Used Gun | 241 (2.4) | 27 (2.7) | 28 (1.9) | 18 (1.9) | 26 (1.1) | 117 (16.2) | 25 (2.2) |
| **Sexual IPV** |  |  |  |  |  |  |  |
| Forced Sex | 1387 (13.6) | 195 (8.2) | 156 (8.8) | 119 (12.2) | 420 (16.6) | 357 (41.8) | 140 (9.7) |
| Forced Sex by Marital Status | 1577 (15.5) | 84 (3.5) | 270 (15.2) | 146 (15.0) | 600 (23.8) | 360 (42.3) | 117 (8.2) |
| **Tertiles of Gender Equitable Attitudes** |  |  |  |  |  |  |  |
| Low Tertile | 3320 (32.6) | 315 (13.2) | 1354 (74.7) | 40 (4.0) | 171 (6.6) | 175 (20.3) | 1265 (82.5) |
| Middle Tertile | 3669 (36.1) | 1332 (55.6) | 418 (23.1) | 183 (18.3) | 1114 (43.2) | 373 (43.2) | 249 (16.2) |
| High Tertile | 3189 (31.3) | 748 (31.2) | 40 2.2) | 775 (77.7) | 1291 (50.1) | 316 (36.6) | 19 (1.2) |

<sup>a</sup>Proportions (column percentages) are shown, may not equal to 100% due to rounding.
<sup>b</sup>M = mean, SD = standard deviation

### Latent Class Solution

Fit statistics indicate that the five-class model was the optimal solution. Adjusted Bayesian information criterion was small for the five-class model. The entropy of 0.81 for the five-class model also suggested an adequate separation of the classes. The Lo-Mendell-Rubin likelihood ratio test p-value of 0.45 for the five-class model indicated that there were no significant improvements made to the overall fit when a sixth class was included.

Overall, the five-class model illustrated distinct and meaningful classes. The *Low Physical/Emotional/Economic/Sexual IPV Perpetration* class (hereafter, Low All Forms of IPV Perpetration) represented 63.1% (n=6406) and was characterized as having a low probability of perpetrating physical, sexual, emotional and economic IPV (**Figure 1**). The *High Physical/Emotional/Economic and Low Sexual IPV Perpetration* class (hereafter, High Physical/Emotional/Economic IPV Perpetration) represented 8.4% (n=851) of the sample and was characterized as high probabilities of perpetrating physical, emotional and economic IPV, and low probabilities of perpetrating sexual IPV. The *High Physical/Sexual/Emotional/Economic IPV Perpetration* class (hereafter, High All Forms of IPV Perpetration) represented 7.6% (n=768) of the sample and was characterized as high probabilities of perpetrating physical, sexual, emotional and economic IPV. The *High Sexual/Emotional and Low Physical/Economic IPV Perpetration* class (hereafter, High Sexual/Emotional IPV Perpetration) represented 7.3% (n=742) of the sample and was characterized as high probabilities of perpetrating sexual and emotional IPV, and low probabilities of perpetuating physical and economic IPV. The *High Emotional and Low Physical/Sexual/Economic IPV Perpetration* class (hereafter, High Emotional IPV Perpetration) represented 13.7% (n=1393) of the sample and was characterized as high probabilities of perpetrating emotional IPV, and low probabilities of perpetrating physical, sexual and economic IPV.

**Figure 1.**
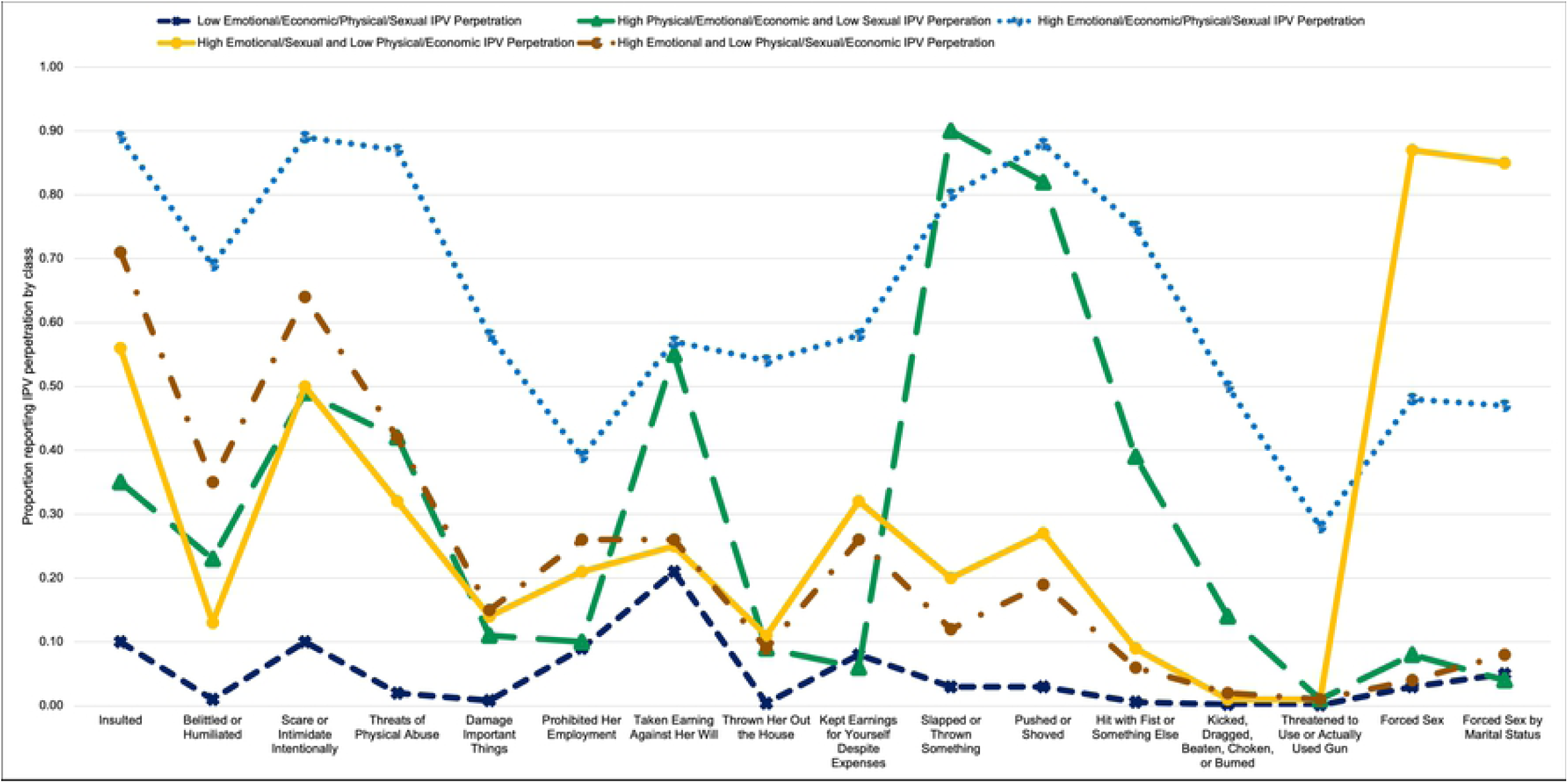
Probability of reporting each IPV perpetration behavior by latent class. 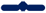 Low Emotional/Economic/Physical/Sexual IPV Perpetration 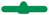 High Physical/Emotional/Economic and Low Sexual IPV Perperation 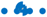 High Emotional/Economic/Physical/Sexual IPV Perpetration 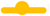 High Emotional/Sexual and Low Physical/Economic IPV Perpetration 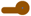 High Emotional and Low Physical/Sexual/Economic IPV Perpetration

### Associations between Gender Equitable Attitudes and Latent Class Membership

There were three significant associations between gender equitable attitudes and latent class membership while adjusting for age, education, employment, married/cohabited and number of children (**Figure 2**). The odds of being assigned to the *Low All Forms of IPV Perpetration* class than the *High All Forms of IPV Perpetration* class was lower for men in the middle tertile group than men in the high tertile group for gender equitable attitudes (OR (95% CI) = .64 (.52, .80), *p*=.00). The odds of being assigned to the *High Emotional IPV Perpetration* class than *the High All Forms of IPV Perpetration* class was greater for men in the low tertile group than men in the high tertile group for gender equitable attitudes (OR (95% CI) = 1.54 (1.06, 2.23), *p*=.02). The odds of being assigned to the *High Physical/Emotional/Economic IPV Perpetration* class than the *High All Forms of IPV Perpetration* class was lower for men in the low tertile group than men in the high tertile group for gender equitable attitudes (OR (95% CI) = .54 (.36, .81), *p*=.003).

**Figure 2a.**
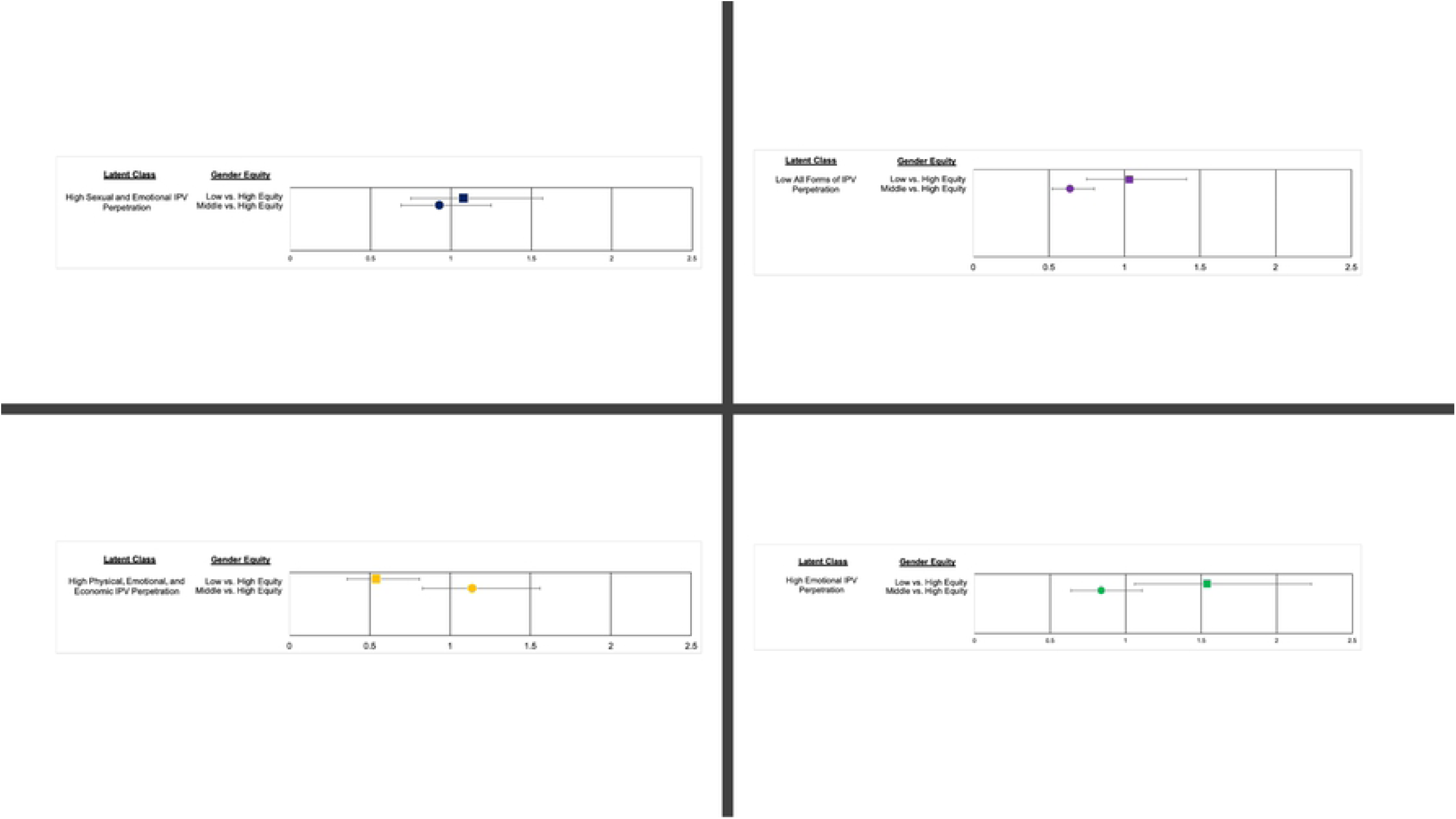
Associations between Latent Classes and Gender Equitable Attitudes among Male Participants Recruited for UN Multi-Country Study on Men and Violence (total sample) *Note*. High All Forms of IPV Perpetration latent class is not shown because it is the reference group. Covariates include age, education, employment, married/cohabitating, whether they had kids and accounted for multistage clustering. <u>Figure Legend</u> *Low vs. High Equity* 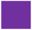 *Middle vs. High Equity* 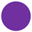 **Figure 2b Associations between Latent Classes and Gender Equitable Attitudes among Male Participants Recruited for UN Multi-Country Study on Men and Violence (total sample)** *Note*. High All Forms of IPV Perpetration latent class is not shown because it is the reference group. Covariates include age, education, employment, married/cohabitating, whether they had kids and accounted for multistage clustering. <u>Figure Legend</u> *Low vs. High Equity* 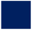 *Middle vs. High Equity* 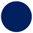 **Figure 2c Associations between Latent Classes and Gender Equitable Attitudes among Male Participants Recruited for UN Multi-Country Study on Men and Violence (total sample)** *Note*. High All Forms of IPV Perpetration latent class is not shown because it is the reference group. Covariates include age, education, employment, married/cohabitating, whether they had kids and accounted for multistage clustering. <u>Figure Legend</u> *Low vs. High Equity* 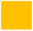 *Middle vs. High Equity* 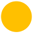 **Figure 2d Associations between Latent Classes and Gender Equitable Attitudes among Male Participants Recruited for UN Multi-Country Study on Men and Violence (total sample)** *Note*. High All Forms of IPV Perpetration latent class is not shown because it is the reference group. Covariates include age, education, employment, married/cohabitating, whether they had kids and accounted for multistage clustering. <u>Figure Legend</u> *Low vs. High Equity* 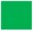 *Middle vs. High Equity* 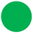

Post-hoc analyses were conducted to explore how relationships between GEM domains (e.g., toxic masculinity, gender roles, sexuality, violence and reproductive health) and latent class membership while adjusting for age, education, employment, married/cohabited and number of children (**Table 2**). A descriptive explanation of post-hoc findings can be found in **Appendix A**.

**Table 2.** Post Hoc Results of Associations between Latent Classes and Domains of Gender Equitable Attitudes among Male Participants Recruited for UN Multi-Country Study on Men and Violence (total sample)

| Class Comparisons | OR (95% CI) | *p* |
| --- | --- | --- |
| **Low All Forms of IPV Perpetration vs. High All Forms of IPV Perpetration** |  |  |
| Gender | 1.23 (.95, 1.60) | .11 |
| Sexuality | 1.01 (.79, 1.28) | .94 |
| Masculinity | 1.13 (1.06, 1.25) | .02 |
| Reproductive Health | .73 (.67, .79) | .00 |
| Violence | 1.69 (1.55, 1.84) | .00 |
| **High Sexual/Emotional IPV Perpetration vs. High All Forms of IPV Perpetration** |  |  |
| Gender | 1.66 (1.16, 2.38) | <.01 |
| Sexuality | 1.05 (.77, 1.44) | .76 |
| Masculinity | .94 (.83, 1.08) | .43 |
| Reproductive Health | .83 (.75, .93) | <.01 |
| Violence | 1.57 (1.39, 1.76) | .00 |
| **High Emotional IPV Perpetration vs. High All Forms of IPV Perpetration** |  |  |
| Gender | 1.54 (1.11, 2.12) | <.01 |
| Sexuality | 1.07 (.79, 1.44) | .66 |
| Masculinity | 1.23 (1.10, 1.38) | .00 |
| Reproductive Health | .67 (.61, .73) | .00 |
| Violence | 1.40 (1.28, 1.54) | .00 |
| **High Physical/Emotional/Economic IPV Perpetration vs. High All Forms of IPV Perpetration** |  |  |
| Gender | .91 (.64, 1.31) | .62 |
| Sexuality | 1.52 (1.10, 2.08) | .01 |
| Masculinity | .97 (.83, 1.13) | .72 |
| Reproductive Health | 1.32 (1.18, 1.47) | .00 |
| Violence | .97 (.83, 1.13) | .70 |

*Note.* Covariates include age, education, employment, married/cohabitating, whether they had kids and accounted for multistage clustering.

## DISCUSSION

To date, empirical IPV research has largely ignored the presence of a potential pattern or lack thereof in men’s IPV perpetration behaviors. This is one of the first studies to examine distinct patterns of IPV perpetration among men in the LMIC context as well as assess gender attitudes and norms as a predictor of those distinct patterns. Using latent class analysis, the present findings indicate five distinct patterns of IPV perpetration among men in the Asia-Pacific region. More than one in three men were characterized in a class indicative of some form of IPV perpetration. Men’s use of IPV has been extensively examined in LMICs,^4,5,18^ however, our study adds to this robust literature by utilizing latent class analysis to identify distinct patterns in the Asia-Pacific region. Further, our study showed significant associations between men’s endorsement of gender equitable attitudes and patterns of IPV perpetration. Our findings suggest that men’s views on gender equity could influence the heterogeneity in men’s use of IPV perpetration. Moreover, distinct domains of gender inequity (e.g., sexuality, toxic masculinity) could provide additional context for some of these associations. Collectively, these findings illustrate ways in which gender inequities contribute to distinct IPV perpetration behaviors exhibited by men in the Asia-Pacific region. Gender-transformative programming and interventions may improve reductions in IPV perpetration among men in the Asia-Pacific region by tailoring intervention components to men’s distinct pattern of IPV perpetration.

Our findings illustrate significant heterogeneity in men’s perpetration of IPV as five classes emerged. For instance, among the four latent classes with a high probability of IPV perpetration, emotional IPV perpetration (e.g., belittling, insults) was the only form of IPV that was present across four latent classes. This finding suggests that emotional IPV is a prevalent form of violence perpetrated by men in the Asia-Pacific region. Emotional IPV also tended to co-occur with other forms of IPV (e.g., physical, sexual), but it is worth noting that one latent class was characterized as men who only used emotional IPV against women (i.e., *High Emotional IPV Perpetration*). Emotional abuse was prevalent among IPV perpetrators, but there was variation in physical, sexual and economic IPV perpetrated by men. For example, high levels of physical IPV and high levels of economic IPV tended to co-occur and this was present in two classes (i.e., *High Physical/Emotional/Economic IPV Perpetration*; and *High All Forms of IPV Perpetration*). Moreover, there was a latent class characterized as high levels of sexual and emotional IPV perpetration, but low levels of physical and economic IPV perpetration (i.e., *High Emotional/Sexual IPV Perpetration*). It is worth noting that some classes of men’s IPV perpetration are consistent with previous research identifying classes of women’s IPV victimization in LMICs. In LMICs, the prevalence of sexual IPV reported by ever-partnered women ranges between 6-59%.^19^ This, in combination with our findings, suggests that men who perpetrate sexual IPV are likely to also use emotional abuse towards their intimate partners. Lastly, the largest latent class was characterized as men who reported little to no IPV perpetration against female intimate partners (i.e., *Low All Forms of IPV Perpetration*). Although the majority of men in this sample were classified in the nonviolent latent class, nearly 37% of men were classified in an IPV perpetration class which is consistent with global estimates of IPV victimization experienced by women.^19^

There were significant associations between men’s endorsement of gender equitable attitudes and their latent class membership. Generally, our findings are consistent with previous literature on gender equitable attitudes,^11,20,21^ suggesting that more gender equitable attitudes relate to less violence being perpetrated by men. For example, men in the low and middle tertiles for gender equitable attitudes had a greater likelihood of being classified in the *High All Forms of IPV Perpetration* class than the *High Physical/Emotional/Economic IPV Perpetration* and *Low All Forms of IPV Perpetration* classes, respectively. These findings suggest that gender equitable attitudes are important contributors of men’s typologies of IPV perpetration. Socially constructed roles between women and men can manifest as power differentials and result in men’s perpetration of violence against women.^20^ Gender-transformative interventions that address how social constructions of masculinity and femininity shape and define gender roles in relationships may help reduce men’s likelihoods of using all types of IPV aging women.^22,23^

It is noteworthy that men in the low tertile for gender equitable attitudes had a greater odds of being classified in the *High Emotional IPV Perpetration* class than the *High All Forms of IPV Perpetration* class. It is possible that this unique finding could be explained by previous research examining how different risk and protective factors are associated with different types of IPV perpetration. For example, a previous study found that men who strongly endorsed sexual cultural scripting norms (e.g., socially constructed sexual roles and expectations between women and men) had a greater likelihood of perpetrating emotional IPV; while men who strongly endorsed traditional masculine norms (e.g., societal views of men’s actions, feelings, behaviors) had a greater likelihood of perpetrating physical IPV.^24^ Building from this previous research, it is possible that men with strong endorsement of sexual cultural scripting norms but weaker endorsement of traditional masculine norms may perpetrate emotional IPV (e.g., belittling) towards their female partner, if she resists these cultural scripting norms (e.g., rejects his sexual advances). It is also possible that men who would be classified in the *High Emotional IPV Perpetration* class have differential access to protective factors (i.e., more social support) that prevent perpetration of physical, sexual and economic IPV than men in the *High All Forms of IPV Perpetration* class. It would be useful for future research to examine differences in protective factors based upon men’s typologies of IPV perpetration in order to inform strengths-based adaptive primary prevention interventions.

Notably, our post-hoc analyses indicated that men with equitable gender attitudes across all domains (i.e., toxic masculinity, gender roles, reproductive health, violence and sexuality) tended to be classified in a typology with fewer types of IPV perpetrated but there were some key differences. Specifically, every domain was not significantly associated with men’s typologies. For example, equitable attitudes in the domains of violence, masculinity and gender did not significantly correlate with men in the *High Physical/Emotional/Economic IPV Perpetration* class vs. the *High All Forms of IPV Perpetration* class. Further, men with more equitable attitudes in the sexuality and reproductive health domains tended to be classified in the *High Physical/Emotional/Economic IPV Perpetration* class than the *High All Forms of IPV Perpetration* class. It is possible that specific domains of gender inequitable attitudes are differentially impacting men’s motivations to perpetrate each IPV type. Additional research is needed to assess which mechanisms, if any, may explicate the differential impacts of gender equitable attitude domains on men’s typology of IPV perpetration. It may also be useful for intervention scientists to examine men’s typology of IPV perpetration as an effect modifier of intervention efficacy. If significant, it may suggest that gender transformative interventions tailor the dose and type of intervention component (e.g., gender equitable domains) to men’s typology of IPV perpetration, such that men in the *High Physical/Emotional/Economic IPV Perpetration* class receive a lower intervention dose in regards to equitable attitudes in the sexuality and reproductive health than men in the *High All Forms of IPV Perpetration* class. These findings also suggest the importance of investigating differential treatment effects by latent class as part of the rapidly growing body of work on IPV prevention in LMICs.

The present study has some limitations. This study is based on the collection of self-reported data, and IPV perpetration may be under-reported, if perceived as a private matter. Under-reporting of IPV perpetration may result in misclassification of latent class membership among this sample, which could result in a lower prevalence across the latent classes and weaken our regression estimates. The study was first conducted in Bangladesh and as a result, the sexual IPV questions were expanded to include coerced sex; therefore, the sexual IPV questions in Bangladesh were different from the questions asked in the other countries, which may have influenced the prevalence. The cross-sectional nature of the data makes it difficult to assess causality. Although we accounted for the clustering effects of the sampling method, this study did not examine these associations by country.

## CONCLUSIONS

Despite these limitations, to our knowledge, this is the first study to use latent class analysis to identify men’s typology of intimate partner violence perpetration and examine its association with gender equitable attitudes. Our findings suggest that IPV perpetration is prevalent among men in the Asia-Pacific region, and five latent classes were identified based on men’s perpetration of IPV. In general, men with more gender equitable attitudes tended to be in a latent class with fewer types of IPV perpetrated than men with less gender equitable attitudes.There is also evidence to suggest that specific domains of gender equitable attitudes may differentially impact men’s typology of IPV perpetration. Findings from this study highlight the importance of continual support for gender transformative interventions addressing IPV perpetration, however an adaptive approach that is personalized to men’s typology of IPV perpetration may have greater impact to reduce violence and promote safety for women in the Asia-Pacific region.

## Data Availability

Additional details about the data and its availability can be accessed through SVRI at http://www.svri.org/what-we-do/research-support/un-multi-country-study-men-and-violence#:~:text=The%20UN%20Multi%2DCountry%20Study,perpetration%20of%20violence%20against%20women.

http://www.svri.org/what-we-do/research-support/un-multi-country-study-men-and-violence

## Contributors

TCW and JG conceived of the study concept and design. MK did the literature searches. TCW did the analysis. TCW, MK, NH and JG agreed with manuscript results and conclusions, and edited the manuscript.

## Acknowledgements

The current study used data collected by the Multi-Country Study on Men and Violence in Asia and the Pacific, coordinated and funded by Partners for Prevention, a UN Development Programme, UN Population Fund (UNFPA), UN Entity for Gender Equality and the Empowerment of Women (UN Women and United Nations Volunteers (UNV) regional joint programme for gender-based violence prevention in Asia and the Pacific. The views expressed in this study are those of the authors and do not necessarily represent those of the UN, including UNDP, UNFPA, UN Women, UNV or UN Member States.

The Sexual Violence Research Initiative (SVRI) has taken over the management of the United Nations Multi-Country Study on Men and Violence in Asia and the Pacific quantitative data set. We are grateful to SVRI for providing our research team with the data necessary to conduct the data analyses and produce the resulting manuscript.

## Data sharing statement

Additional details about the data and its availability can be accessed through SVRI at http://www.svri.org/what-we-do/research-support/un-multi-country-study-men-and-iolence#:~:text=The%20UN%20Multi%2DCountry%20Study,perpetration%20of%20violence%20against%20women.

## Details of ethics approval

Study procedures have been approved by the [Institution masked for peer review] IRB (Protocol #).

## References

1. Kapiga S, Harvey S, Mshana G, et al. A social empowerment intervention to prevent intimate partner violence against women in a microfinance scheme in Tanzania: findings from the MAISHA cluster randomised controlled trial. The Lancet Global Health 2019; 7(10): e1423–e34.

2. Pronyk PM, Hargreaves JR, Kim JC, et al. Effect of a structural intervention for the prevention of intimate-partner violence and HIV in rural South Africa: a cluster randomised trial. The lancet 2006; 368(9551): 1973–83.

3. Gupta J, Falb KL, Lehmann H, et al. Gender norms and economic empowerment intervention to reduce intimate partner violence against women in rural Côte d’Ivoire: a randomized controlled pilot study. BMC Int Health Hum Rights 2013; 13(1): 46.

4. Gibbs A, Jacobson J, Kerr Wilson A. A global comprehensive review of economic interventions to prevent intimate partner violence and HIV risk behaviours. Global health action 2017; 10(sup2): 1290427.

5. Fulu E, Jewkes R, Roselli T, Garcia-Moreno C. Prevalence of and factors associated with male perpetration of intimate partner violence: findings from the UN Multi-country Cross-sectional Study on Men and Violence in Asia and the Pacific. The lancet global health 2013; 1(4): e187–e207.

6. Jewkes R, Fulu E, Roselli T, Garcia-Moreno C. Prevalence of and factors associated with non-partner rape perpetration: findings from the UN Multi-country Cross-sectional Study on Men and Violence in Asia and the Pacific. The lancet global health 2013; 1(4): e208–e18.

7. Fulu E, Miedema S, Roselli T, et al. Pathways between childhood trauma, intimate partner violence, and harsh parenting: findings from the UN Multi-country Study on Men and Violence in Asia and the Pacific. The Lancet Global Health 2017; 5(5): e512–e22.

8. World Health Organization. Putting women first: ethical and safety recommendations for research on domestic violence against women: Geneva: World Health Organization, 2001.

9. Garcia-Moreno C, Jansen HA, Ellsberg M, Heise L, Watts CH. Prevalence of intimate partner violence: findings from the WHO multi-country study on women’s health and domestic violence. The Lancet 2006; 368(9543): 1260–9.

10. Jewkes R, Sikweyiya Y, Morrell R, Dunkle K. Gender inequitable masculinity and sexual entitlement in rape perpetration South Africa: findings of a cross-sectional study. PLoS One 2011; 6(12): e29590.

11. Pulerwitz J, Barker G. Measuring attitudes toward gender norms among young men in Brazil: development and psychometric evaluation of the GEM scale. Men and Masculinities 2008; 10(3): 322–38.

12. Shattuck D, Burke H, Ramirez C, et al. Using the Inequitable Gender Norms scale and associated HIV risk behaviors among men at high risk for HIV in Ghana and Tanzania. Men and Masculinities 2013; 16(5): 540–59.

13. Lusey H, San Sebastian M, Christianson M, Edin KE. Factors associated with gender equality among church-going young men in Kinshasa, Democratic Republic of Congo: a cross-sectional study. International journal for equity in health 2017; 16(1): 213.

14. Vu L, Pulerwitz J, Burnett-Zieman B, Banura C, Okal J, Yam E. Inequitable gender norms from early adolescence to young adulthood in Uganda: Tool validation and differences across age groups. J Adolesc Health 2017; 60(2): S15–S21.

15. McCutcheon AL. Latent class analysis: Sage; 1987.

16. Celeux G, Soromenho G. An entropy criterion for assessing the number of clusters in a mixture model. Journal of classification 1996; 13(2): 195–212.

17. Muthén L, Muthén B. Mplus statistical modeling software: Release 7.0. Los Angeles, CA: Muthén & Muthén 2012.

18. Mulawa MI, Kajula LJ, Maman S. Peer network influence on intimate partner violence perpetration among urban Tanzanian men. Cult Health Sex 2018; 20(4): 474–88.

19. Devries KM, Mak JY, Garcia-Moreno C, et al. The global prevalence of intimate partner violence against women. Science 2013; 340(6140): 1527–8.

20. Chirwa ED, Sikweyiya Y, Addo-Lartey AA, et al. Prevalence and risk factors of physical or sexual intimate violence perpetration amongst men in four districts in the central region of Ghana: Baseline findings from a cluster randomised controlled trial. PLoS One 2018; 13(3): e0191663.

21. McCarthy KJ, Mehta R, Haberland NA. Gender, power, and violence: A systematic review of measures and their association with male perpetration of IPV. PLoS One 2018; 13(11): e0207091.

22. Dworkin SL, Fleming PJ, Colvin CJ. The promises and limitations of gender-transformative health programming with men: critical reflections from the field. Cult Health Sex 2015; 17(sup2): 128–43.

23. Dworkin SL, Treves-Kagan S, Lippman SA. Gender-transformative interventions to reduce HIV risks and violence with heterosexually-active men: a review of the global evidence. AIDS Behav 2013; 17(9): 2845–63.

24. Willie TC, Khondkaryan E, Callands T, Kershaw T. “Think Like a Man”: How Sexual Cultural Scripting and Masculinity Influence Changes in Men’s Use of Intimate Partner Violence. Am J Community Psychol 2018.

